# A control approach to the Covid-19 disease using a SEIHRD dynamical model

**DOI:** 10.1101/2020.05.27.20115295

**Authors:** Fernando Pazos, Flavia E. Felicioni

## Abstract

The recent worldwide epidemic of Covid-19 disease, for which there is no vaccine or medications to prevent or cure it, led to the adoption of public health measures by governments and populations in most of the affected countries to avoid the contagion and its spread. These measures are known as nonpharmaceutical interventions (NPIs) and their implementation clearly produces social unrest as well as greatly affects the economy. Frequently, NPIs are implemented with an intensity quantified in an ad hoc manner. Control theory offers a worthwhile tool for determining the optimal intensity of the NPIs in order to avoid the collapse of the healthcare system while keeping them as low as possible, yielding in a policymakers concrete guidance. We propose here the use of a simple proportional controller that is robust to large parametric uncertainties in the model used.

## 1 Introduction

The novel SARS-CoV-2 Coronavirus, which produces the disease known as Covid-19, was first reported on December 2019 in Wuhan, province of Hubei, China. With amazing speed it spread to the majority of the countries in the world. The outbreak has been declared as a public health emergency of international concern by the World Health Organization (WHO) on Jan. 30, 2020 and as a pandemic on March 11.

At the moment, there is no vaccine against this virus or effective medicines to cure the disease. Health systems only try to mitigate its consequences to avoid complications and fatal outcomes. This disease showed a great capacity of contagion and high fatality rates (see updated reports in [WM2]).

Patients affected by this disease experience a number of symptoms, not all clearly identified at the moment, but which are mainly cough, breathing difficulties, fever, loss of taste and smell and extreme tiredness. Frequently, patients develop a form of viral pneumonia that requires hospitalization and artificial mechanical ventilation in intensive care units. The large number of patients affected by this disease threatens to collapse public health systems, increasing the fatality rates by lack of available health assistance.

In this context, is very important to predict the trend of the epidemic in order to plan effective strategies to avoid its spread and to determine its impact. As the contagion is produced very easily by simple contact between people, several measures were adopted by the governments, public health systems and populations in order to reduce the transmission by reducing contact rates. Examples of these measures, the so called nonpharmaceutical interventions (NPIs) adopted during this period include the closing of schools, churches, bars, factories, quarantine or physical-distancing policies, confinement of people in their homes, lockdown, among other social impositions that produce discomfort and clearly harm the economy.

This goal sparked many articles and studies recently published on the epidemic behavior. A number of them are addressed towards determining a mathematical model that represents the dynamics of different agents involved in a population affected by the disease. The dynamic described by the model aims to make possible to answer crucial issues, such as the maximum number of individuals that will be affected by the disease and when that maximum will occur, and makes key predictions concerning the outbreak and eventual recovery from the epidemic. This information allows to devise public policies and strategies to mitigate the social impact and reduce the fatality rate. The seminal work [FLNG^+^20] exemplifies and analyses different strategies to control the transmission of the virus.

Most of the models adopted to represent the dynamical behavior of the Covid-19 are based on the SIR model (see [Abd20] and references therein). The SIR model is a basic representation widely used which describes key epidemiological phenomena. It assumes that the epidemic affects a constant population of *N* individuals. The model neglects demography, i.e. births and deaths by other causes unrelated to the disease^1^.

The population is broken into three non-overlapping groups corresponding to stages of the disease:

- **Susceptible (S).** The population susceptible to acquire the disease.
- **Infected (I).** The population that has acquired the virus and can infect others.
- **Recovered (R).** The population that has recovered from infection and presumed to be no longer susceptible to the disease^2^.

A brief description of these compartments is given below. Susceptible people are those who have no immunity and they are not infected. An individual in group S can move to group I by infection produced through contact with an infected individual. Group I are people who can spread the disease to susceptible people. Finally, an infected individual recovered from the disease is moved from the group I to the group R.Some references (see, for example [LZG^+^20,SSB20]) considers the group R as removed population, or closed cases, which includes those who are no longer infectious from recovery and the ones who died from the disease. The summation of these three compartments in the SIR model remains constant and equals the initial number of population *N*.

In order to describe better the spread of epidemics, many works (see, for example [Kan20,GV20,SHD20,Nes20]) adopted the SEIR model. In the SEIR model a fourth group denoted as Exposed (E) is added between the group S and the group I:

- **Exposed (E).**The population that has been infected with the virus, but not yet in an infective stage capable of transmitting the virus to others.

This compartment is dedicated to those people who are infectious but they do not infect others for a period of time namely incubation or latent period.

Other works (see [Abd20] for example) consider an additional compartment at the end of the SIR or of the SEIR model to distinguish between recovered and death cases:

- **Dead (D).**The population dead due to the disease.

Thus, these models become the SIRD or the SEIRD models, respectively.

Other works as [TBT^+^20,LZG^+^20,SSB20] consider the existence of other groups seeking to match the models proposed with the numbers obtained from the actual Covid-19 disease.

The work presented in [GBB^+^20] deserves to be mentioned. This work studies the evolution of the Covid-19 in Italy, and proposes a model denoted as SIDARTHE, where the letters correspond to eight groups denoted as Susceptible, Infected, Diagnosed, Ailing, Recognized, Threatened, Healed and Extinct respectively. All of them are subgroups of those presented in the SEIR model. This model discriminates between detected and undetected cases of infection, either asymptomatic or symptomatic, and also between different severity of illness, having a group for moderate or mild cases and another one for critical cases that require hospitalization in intensity care units. The authors affirm that the distinction between diagnosed and non-diagnosed is important because nondiagnosed individuals are more likely to spread the infection than diagnosed ones, since the later are typically isolated, and can explain misperceptions of the case fatality rate and the seriousness of the epidemic phenomena. The fact of considering more groups in the SIDARTHE model than in the SEIR model allows better discrimination between the different agents involved in the epidemic evolution, as well as a better differentiation of the role played by each one. However, the fact of considering more groups implies the knowledge of more rates, probabilities and constants that determine the dynamics between the groups. Many of these values are difficult to know in practice, as well as to estimate the population of some groups, such as Ailing (symptomatic infected undetected). The authors choose these constants and quantities to match the model to the actual data. Of course, in order to achieve the goal of better determining public policies, we believe that the existence of some of these groups in the model used is not necessary.

In order to better guide the determination of public policies to mitigate the spread of the virus we propose the use of the control theory.

Control theory has been successfully implemented in several areas other than physical systems control, for which it was initially designed. For example in economics, ecological and biological systems, many works demonstrate the success of its implementation. Of course, regardless of the area focused, a good control strategy depends on the adequate modeling of the dynamical system to be controlled.

The proposal to use control in this epidemic is not new. It has been first presented in [SHD20]. In this work, the authors use the SEIR model to show that a simple feedback law can manage the response to the pandemic for maximum survival while containing the damage to the economy. However, the authors illustrate with several examples the benefits of using feedback control, but they do not present the mathematical control laws as well as they do not prove the convergence of the trajectories in the closed loop system. Examples are implemented by mean of several computational experiments which illustrate the different strategies proposed.

We propose here the use of a simple proportional controller, a standard tool in control theory, to calculate the control action. This variable guides how to determine NPIs in order to avoid the collapse of the health system while reducing the damage to the society and the economy that NPIs inevitably produce.

## 2 The SEIHRD model

This section is addressed to model adequately the disease. A suitable model should avoid making unnecessary classifications in order to obtain key data on the behavior of the epidemic. These data include number of deaths, maximum number of infected people, time at which the maximum infection rate will occur, among other information useful to prevent and reduce the damage produced by the outbreak.

The SEIR model assumes that exposed people have been infected but are not able to transmit the virus before a latency period. We will consider that those people continue to be in the susceptible group S, whereas we consider the group E as people who have been infected but have no symptoms yet and are capable of transmitting the virus. Of course, part of this group will present symptoms after an incubation time (moving to the group I) and another part will remain asymptomatic. Asymptomatic people who have been diagnosed as positive also will be considered in the group I, so this group includes all known positive cases, symptomatic or not.

In addition, a critical issue is the number of infected people who need hospitalization, because the public policies must try to keep this number lower than the capacity of the health care system in order to avoid its collapse. Thus we define an extra group:

- **Hospitalized (H).** The infected population who need hospitalization.

In the group H we do not differentiate between people hospitalized in mild condition and those in intensive care units (ICU_s_), despite the fact that the number of people in the last subgroup is a critical problem due to an even more limited capacity in ICU_s_.

We also consider the population number *N* as a constant, as the SEIR model does.

The progression of this epidemic can be modeled by the rate processes described in Fig. 1.

**Figure 1:**
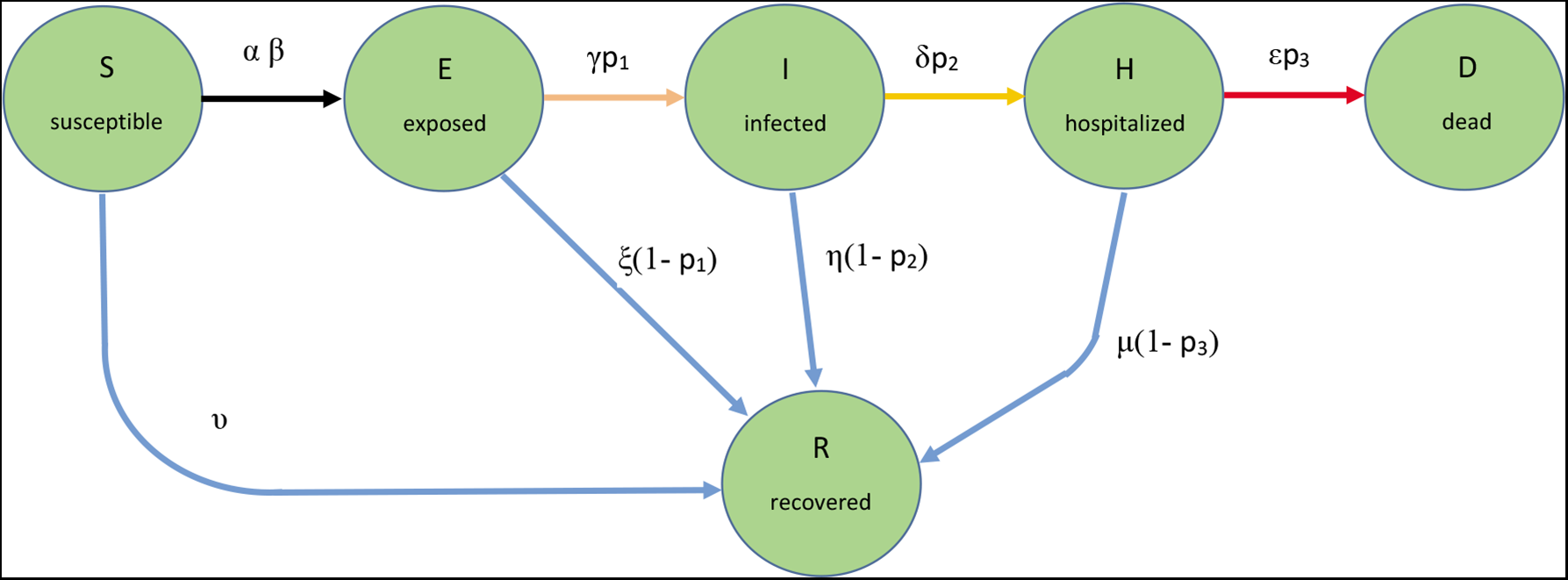
Rate processes that describes the progress between the groups in the SEIHRD model.

The proposed SEIHRD model for the spread of the Covid-19 disease in an uniform population is given by the following deterministic equations, which are presented normalized with respect to the total population *N*.

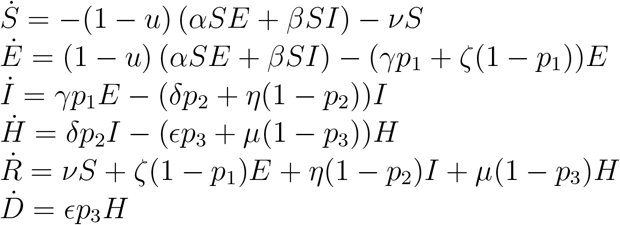

The groups *S, E, I, R, H* and *D* are the state variables of the dynamical system (1). They are always nonnegative. The time derivatives *Ṙ* and *Ḋ* are also nonnegative, because recovered people and death cannot decrease, whereas *Ṡ* is always nonpositive, because we consider that recovered people cannot be reinfected. This fact is represented in Fig. 1 because the states *R* and *D* only have input arrows and the state *S* only has an output arrow. The model (1) is a nonlinear system normalized with respect the population *N*, considered as a constant.Hence *S* + *E* + *I* + *H* +*R* + *D* = 1 and *Ṡ* + *Ė* + *İ* + *Ḣ* + *Ṙ* + *Ḋ* = 0. The system (1) presents an equilibrium point in 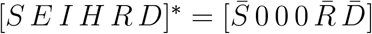, where 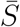 and 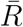 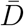 are positive values.

The rate processes are modeled as follows.

- αSE and *βSI* are the transmission rates of the virus between the susceptible and the exposed population (respectively, infected population). *α* and *β* are the probability of disease transmission in a single contact with exposed (infected) people times the average daily number of contacts per person and have units of 1*/day*. Typically, *α* is grater than *β*, assuming that people tend to avoid contact with subjects showing symptoms or diagnosed as positive. Contacts between susceptible people and hospitalized people are neglected, excepting for healthcare workers. The probability of contagion from dead people is also neglected, despite the fact that some cases were recently reported. Of course, recovered people are no longer able to transmit the virus.
- *u*∈[0, 1] is the effectiveness of nonpharmaceutical public health interventions (NPIs). *u*=0 means no intervention and the epidemic grows completely free, whereas *u*=1 implies total elimination of the disease spread.
- ν is the vaccination rate, at which susceptible people became unable to be infected. Unfortunately, in the Covid-19 case *v*=0 yet.
- *p*_1_ is the probability that exposed people develop symptoms,*γ*^−1^ is the average period to develop symptoms, and *ζ*−^1^ is the average time to overcome the disease staying asymptomatic.
- *p*_2_ is the probability that infected people with symptoms require hospitalization,*δ*^−1^ is the average time between infection and the need for hospitalization, and *η*^−1^ is the average time in that infected people recover without hospitalization.
- *p*_3_ is the probability of hospitalized people die, ∊^−1^ is the average time between the hospitalization and the death, and *µ*^−1^ is the average time to recover after hospitalization.

The parameters used in (1) are not very precisely determined and even differ greatly in the literature consulted (see [FLNG^+^20, LGWSR20, Kan20, LZG^+^20, GBB^+^20, Jon19, Org] among many other references). Most of the model adopted in the references adjust these parameters to match real data from different countries. It must be taken into account that some of these parameters, mainly *α* and *β*, are not independent either from the populations and their general health status or their actions.

The parameters *α* and *β* are related with the basic reproduction number *R*_0_, defined as the expected number of secondary cases produced by a single (typical) infection in a completely susceptible population [Jon19].*R*_0_ is not a fixed number, depending as it does on such factors as the density of a community, the general health of its populace, or its medical infrastructure [SHD20]. This is the most important parameter to understand the spread of an epidemic. If *R*_0_ > 1, the epidemic growths and the number of infected people increases. If *R*_0_ < 1, the epidemic decreases and after a certain time disappears, when a large enough number of people acquire antibodies and the so-called herd immunity occurs.

In the actual Covid-19 disease, *R*_0_ was determined to be 2.6 in Wuhan, China [SHD20] (between 2.2 and 2.7 according to [SLX^+^20]), ranging from 2.76 to 3.25 in Italy [SHD20] and even close to 3.28 [LGWSR20]. An important remark is that many works consider *R*_0_ depending on the NPIs, admitting that these actions tend to reduce this number because the contact rates between people decrease. Note that NPIs always occur even in countries where no government action has been taken, because people spontaneously tend to stay at home and to avoid contact with others. This fact explains the disparity of this number in different countries and reported in the references (see [LGWSR20]).

Here, we consider *R*_0_ as a constant reproduction number in the absence of any external action, i.e., as if the disease could spread completely free, which, of course, is an unrealistic scenario. Specifically, the relation between the rates *α* and *β* and *R*_0_, can be calculated in model (1) as in [Jon19,GBB^+^20], resulting

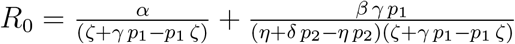

In section 3 we propose a pair of values for parameters *α* and *β* to evaluate different epidemic scenarios.

The effectiveness of the NPIs is considered in the variable *u*, which determines the rate at which susceptible people become exposed.

Several works [LZG^+^20,TBT^+^20,GV20,SSB20] consider these parameters as time dependent, because incorporate in these parameters the impact of governmental actions among other NPIs.

The incubation period is estimated as *γ*^−1^ =5.1 days [Kan20,FLNG^+^20, TWL^+^20].

The probability of developing symptoms *p*_1_ will be roughly estimated as

50% [MKZC20,FLNG^+^20]^3^.

The period to overcome the disease without presenting symptoms is *ζ*^−1^ =14.7 days (deduced from [GBB^+^20]).

The infectious period with no need of hospitalization is widely accepted as 14 days, so *η* = 1*/*14.

The probability to need hospitalization after the infection is *p*_2_ = 19% [Low20,LZG^+^20,SSB20], and the time from symptom onset to hospitalization is *δ*^−1^= 5.5 days [SLX^+^20].

The probability to die after hospitalization is *p*_3_ = 15% according to [WM2,FLNG^+^20], and the average time to die is *∊*^−1^ = 11.2 days [SLX^+^20].

The average time to recovery after hospitalization is *µ*^−1^ = 16 days [FLNG^+^20].

Finally, as it was noted above, there is no vaccine against this disease, so *ν* = 0.

### 2.1 Remark

*Of course, most of these parameters are subject to large inaccuracies, and they differ greatly in the literature consulted. However, as we will show below, the proposed control method is robust for such uncertainties as well as for measurements errors characterized as unreported or undiagnosed cases and inaccuracies in the group quantities*.

## 3 The control strategy

We propose the use of control theory to determine public nonpharmaceuticals interventions (NPIs) in order to control the evolution of the epidemic, avoiding the collapse of health care systems while minimizing harmful effects on the population and on the economy.

As noted in [SHD20], “a properly designed feedback-based policy that takes into account both dynamics and uncertainty can deliver a stable result while keeping the hospitalization rate within a desired approximate range. Furthermore, keeping the rate within such a range for a prolonged period allows a society to slowly and safely increase the percentage of people who have some sort of antibodies to the disease because they have either suffered it or they have been vaccinated, preferably the latter”.

The action law is given by the control variable *u* in (1). No intervention from the public health agencies means *u* = 0, and the disease evolves naturally without control. At the other hand *u* = 1 means the total impossibility of transmitting the virus, which, of course, is an unrealistic scenario.

There are several possible choices of the reference signal or set point of the control system. One of them may be a small enough number of hospitalized people to not affect the capacity of the intensive care units (ICU_s_)available in the health care system. This reference signal maybe nonconstant, it can can go up because of an increase in available beds due to capacity additions in the health care system, by creation of provisory field hospitals, among other similar measures. By other hand, we must bear in mind that the quantities of each group described in (1) are subject to large inaccuracies, due to unreported or undiagnosed cases, except for the number of people diagnosed as positive (I), which is quite well known, the number of hospitalized people (H) and, of course, the number of deaths (D). For that reason the output variable to be fed back only can be the infected population I or the hospitalized population H.

Hence, the goal of the control action is to keep the number of hospitalized people lower than the set point minimizing the external intervention which produces social discomfort and clearly harm the economy.

Therefore, the control action should aim to solve the following constrained optimization problem:

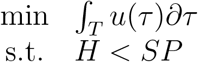

where *T* is a considered period and *SP* is the reference signal (or set point, in the case it is considered as a constant).

As a reference, the World Health Organization recommends a number of 80 hospital beds per 10000 population, which means an index of 0.008, or

0.8%. This number will be used as the SP of the closed loop control system.

However, we must bear in mind that NPIs impact on physical contacts between susceptible and infected or exposed people. If an individual is already infected, hospitalization will be required after at most *δ*^−1^ = 5.5 days or after *δ*^−1^+ *γ*^−1^= 10.6 days on average if the infection was recent. Hence, there exists a delay between the adoption of NPIs and their consequences on hospitalization of people. If the control action if calculated based only on the number of hospitalized people, the following 10.6 days too many people may require hospitalization, exceeding the capacity for medical care. In control jargoon, it means that there are almost two weeks with the system operating in an open loop. Therefore, the control action needs to be calculated as a function of the number of infected people I (the number of exposed people E is quite unknown) in order to avoid future hospitalization requirements in the next 10.6 days at most. This strategy is known as *predictive control*.

Fig. 2 shows the closed loop control system. The variable process is the infected population *I* and the control signal is the effectiveness of the NPI scalar signal *u*.

**Figure 2:**
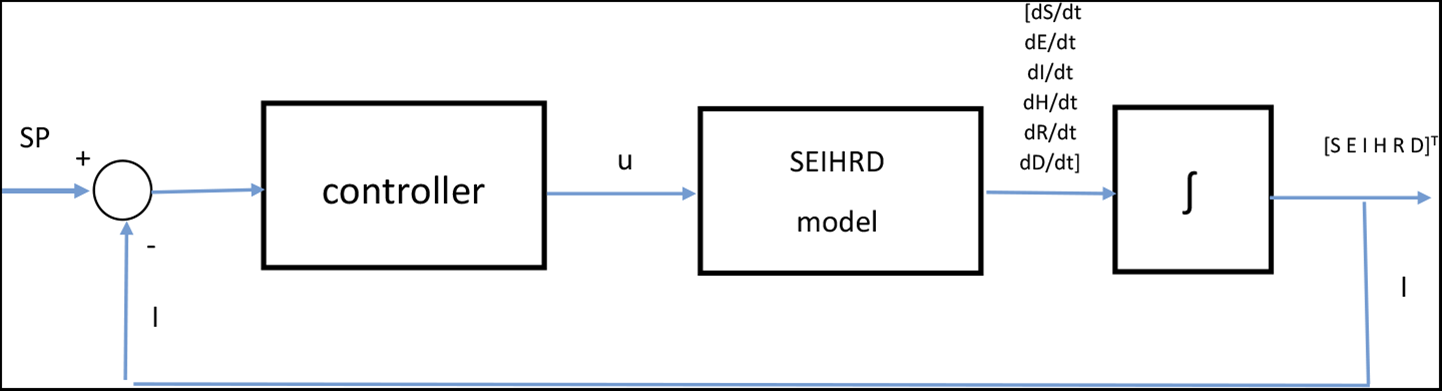
Block diagram of the closed loop control system.

Of course, in practical situations it is necessary to determine which actions and at what level correspond to a certain effectiveness of NPIs, but this issue is outside the scope of this paper.

Next, we show the results of different strategies of NPIs applied on the SEIHRD model.

## 3.1 An open loop control system

In this first series of experiments, we apply a constant control action *u*, that is, the system shown in Fig. 2 is an open loop control one.

We consider as initial conditions *I* = *E* = 0.001, *H* = *R* = *D* = 0, so *S* = 0.998, that is, 0.1% of the population is diagnosed as positive the first day and 0.1% of the population is asymptomatic infected.

During the first days of the epidemic, it was logical to consider that both exposed and infected people could spread the virus at the same ratio because the contagion between humans was not known. Then, this disease could spread in a completely free scenario, in which no action is taken. This scenario has been called “*naif*” by several authors [SSB20,LZG^+^20].

Using the expression (2) with *R*_0_ = 2.8 as in [SSB20,LZG^+^20], and assuming that no actions are taken during the epidemic, then *α* = *β* = 0.1786. The evolution of exposed, infected, hospitalized and dead people in this case is shown in the Fig. 3.

**Figure 3:**
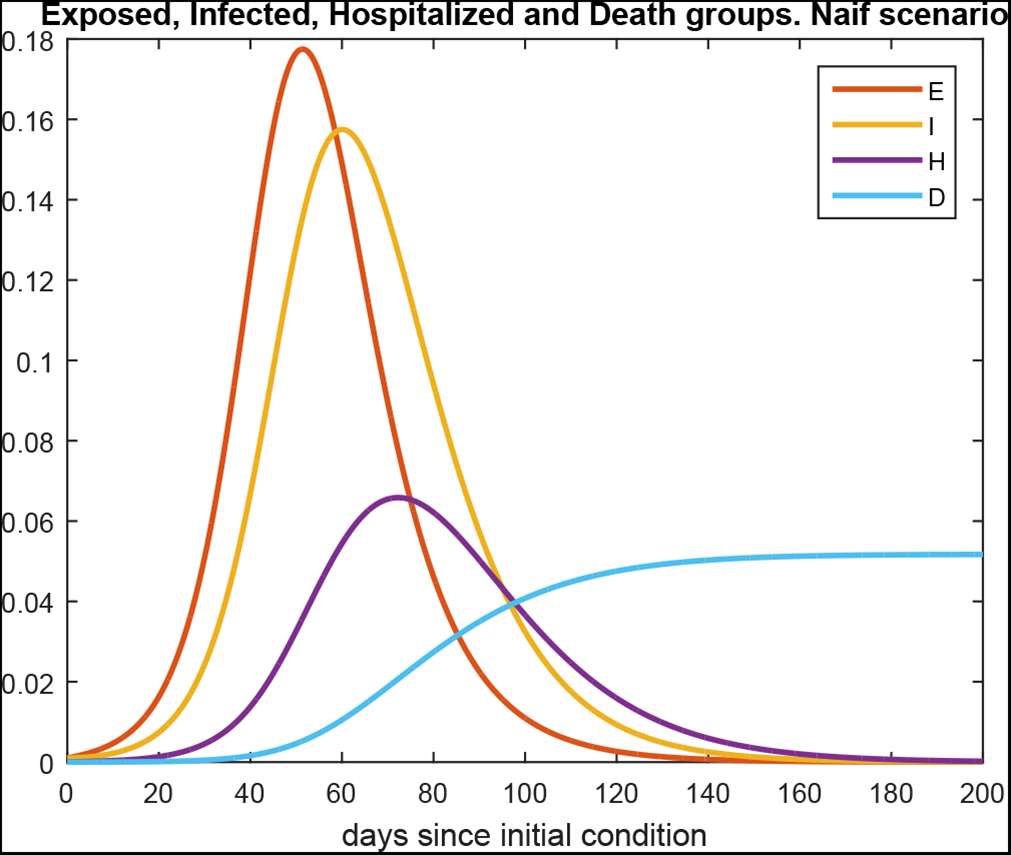
Population of Exposed, Infected, Hospitalized and Deaths group with no NPI. Naif Scenario

In this “*naif*” scenario, and using as initial condition 1 infected and 1 exposed person for different population values (*N >* 1, 000), the maximum are always 17.75% for the exposed and 15.75% for the infected, and the times when these maximums are reached depend on the population value *N* as is shown in the Fig. 4. The delay between both maximums is a constant value of 9 days. Additionally, the number of dead people forecasted by this model is about 5.17% of the total population.

**Figure 4:**
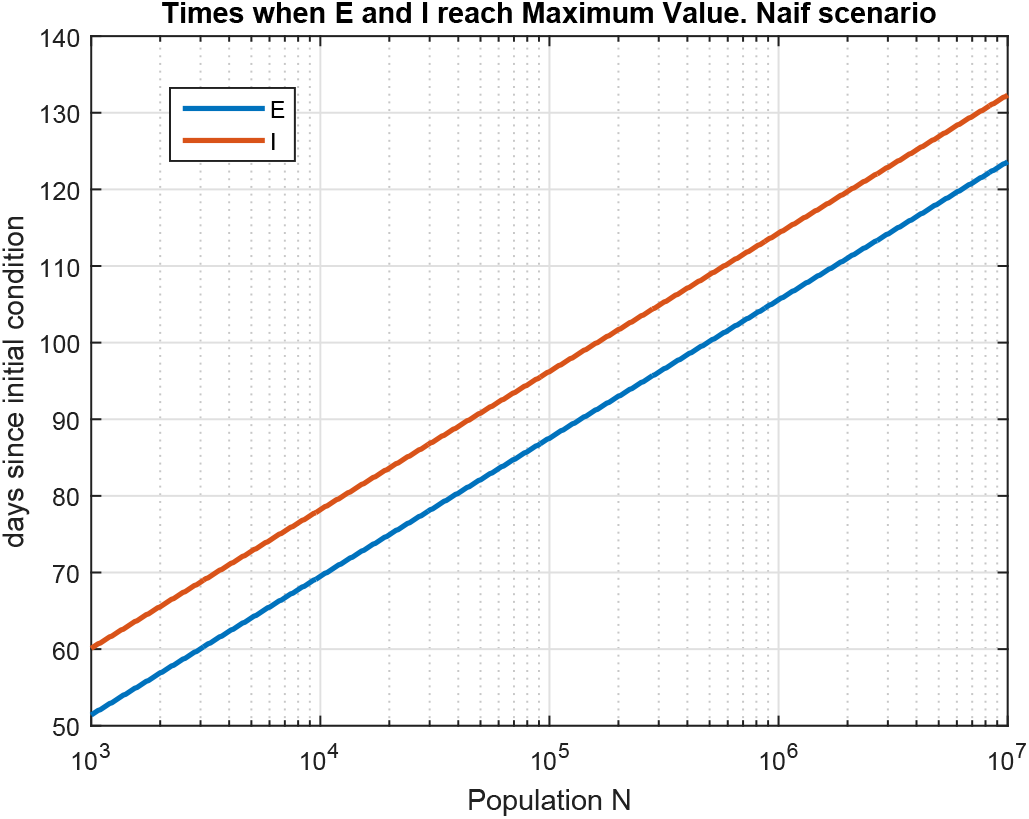
Times when Exposed and Infected gropus reach Maximum Values. Naif Scenario

Clearly, this “*naif*” scenario seemed to be unrealistic since people tend to avoid contact with subjects showing symptoms or diagnosed as positive due to the severity of the Covid-19 disease. In consequence, as we stated before, in a more realistic scenario *α* is greater than *β*. In the rest of this paper we consider *β* = *α/*2 to take into account this assumption.

Fig. 5 shows the areas of every group along the time in the case with no NPI actions for illustrative purposes (with *β* = *α/*2).

**Figure 5:**
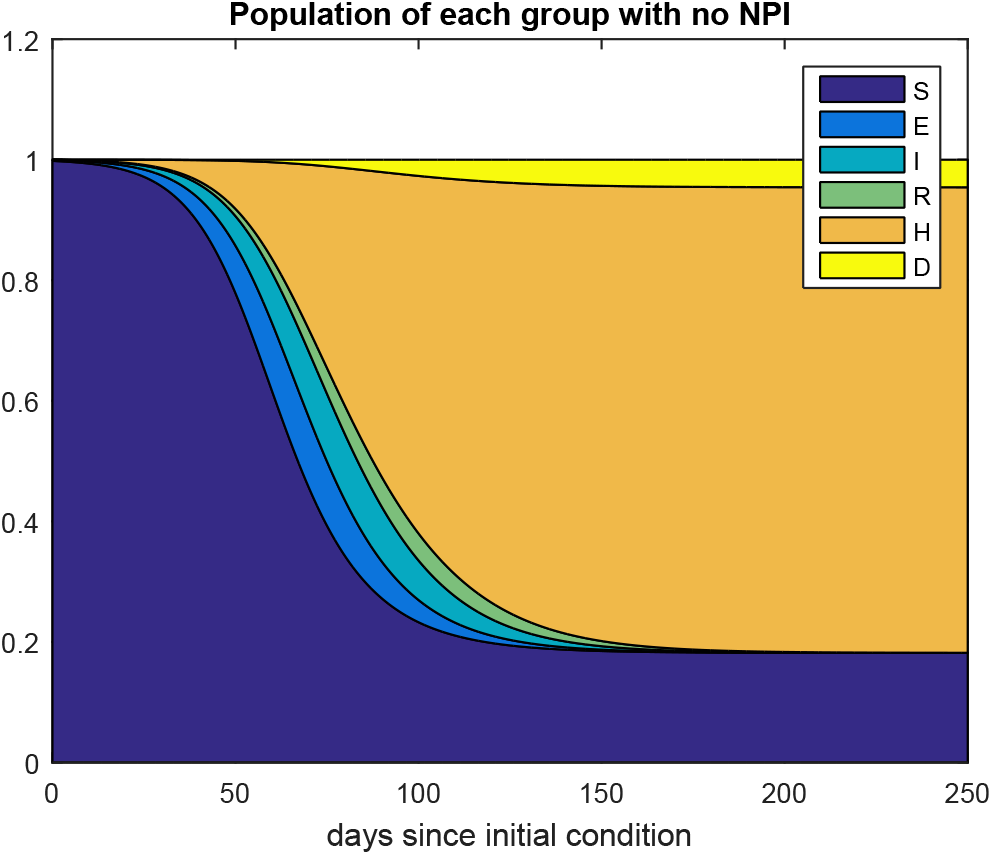
Population of each group with no NPI. *β* = *α/*2

Fig. 6 shows the evolution of the Hospitalized with different constant NPIs effectiveness *u* and the proposed SP.

**Figure 6:**
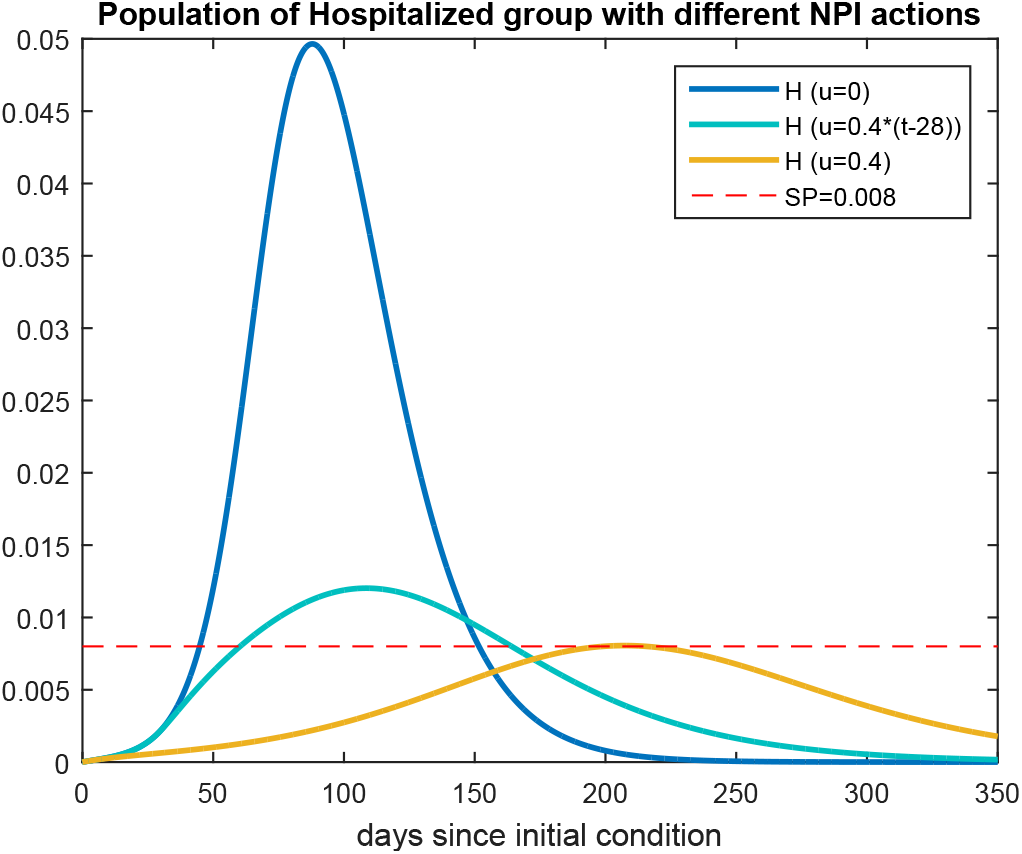
Population of Hospitalized group with no NPI (blue), with a NPI of 40% effectiveness (yellow), with an intervention of 40% effectiveness applied 4 weeks after the appearance of the first case (light blue) and SP (red). *β* = *α/*2

Table 1 reports some results extracted from these simulations.

**Table 1:** Main results of several constants NPI on the SEIHRD model after 250 days.

| | $u = 0$ | $u = 0.4$ | $u = 0.4 \text{ step}(t - 28)$ |
| --- | --- | --- | --- |
| final rate of deaths | 0.0457 | 0.01425 | 0.022 |
| maximum rate of hospitalized | 0.0496 | 0.00801 | 0.0120 |
| area under the curve $u$ vs. $t$ | 0 | 100 | 88.8 |

The results presented in Table 1 show that, if no mitigation policy is adopted (*u* = 0) approximately 81% of the population will be infected and 4.57% will die. On the other hand, a relatively little aggressive NPI, only 40% of effectiveness, is efficient in reducing the final number of deaths as well as the maximum number of hospitalized people, which is a crucial issue in order not to collapse the health system (the maximum value of H reaches the *SP*). Moreover, a late application of this strategy, after 4 weeks since the first case arose, also significantly reduces these numbers.

## 3.2 A proportional controller

In this section, we simulate the behavior of the trajectories described by the normalized system (1) subject to a proportional control action. The objective of the control action is that the number of hospitalized people does not exceed the number of available beds. Of course, this number is highly variable in different countries, and can be increased during the duration of the epidemic with the construction of field hospitals, among other resources.

On the other hand, as noted in Sec. 3, to adopt as feedback variable the number of hospitalized people may lead to an overload of the health system in the following 10.6 days, for which a predictive control must be used that consider the number of infected people I. Not all infected people need hospitalization. Most of the symptomatic cases are mild and remain mild in severity [Low20, SLX^+^20] (1 *− p*_2_ = 81%). So we consider that *p*_2_ = 19% of infected people will need hospitalization in the following *δ*^−^^1^ = 5.5 days.

This number plus the number of people already hospitalized H must remain below the set point. Of course, we neglect the number of beds occupied by patients hospitalized for other diseases.

Hence, the proportional control variable is chosen as

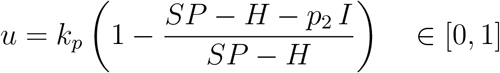

where *k_p_* is the proportional scalar gain with values between [0, 1]. Note that if *I* = 0, *u* = 0, and there is no need of a public intervention because no one is going to require hospitalization on the following 5.5 days, and with *k_p_* = 1, if a percentage of 19% of the infected people is equal to the number of available beds *SP − H, u* = 1 which means that the public intervention must completely avoid the transmission of the virus because all these people will require hospitalization after *δ*^−1^ = 5.5 days on average. Another point of view is to consider that this is a tracking trajectory problem, with a time dependent reference signal equal to *r*(*t*) = *SP − H*(*t*).

We consider the same initial condition than in the former series of experiments, *I* = *E* = 0.001, i.e. 0.1% of the population infected and presenting symptoms at the first day and we suppose that 0.1% of the population is infected and asymptomatic.

Fig. 7 shows the trajectories of the variables vs. time with a gain *k_p_* = 1. Note that the number of people hospitalized is always smaller than the set point.

**Figure 7:**
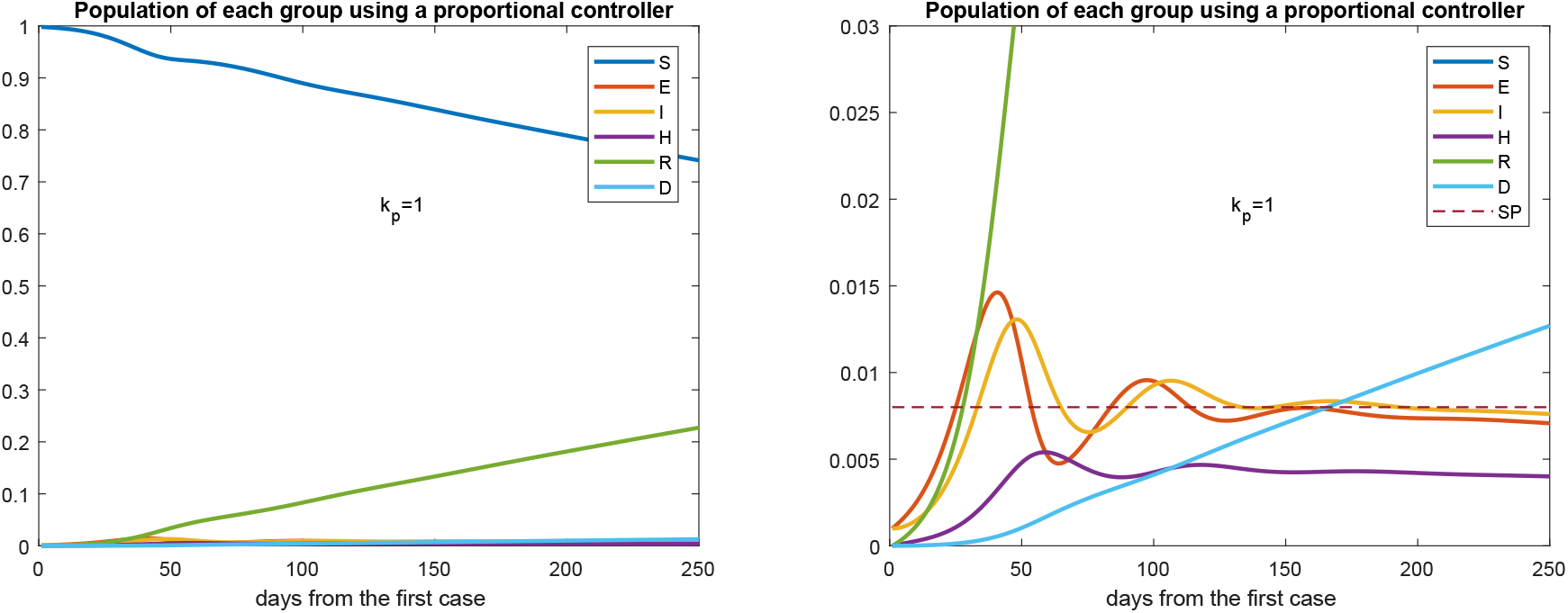
Evolution of every group over time with a proportional control action with gain *k_p_* = 1 (left). The picture at the right is a zoom of that at the left. Set point equal to 0.008.

Fig. 8 shows the control signal vs. time.

**Figure 8:**
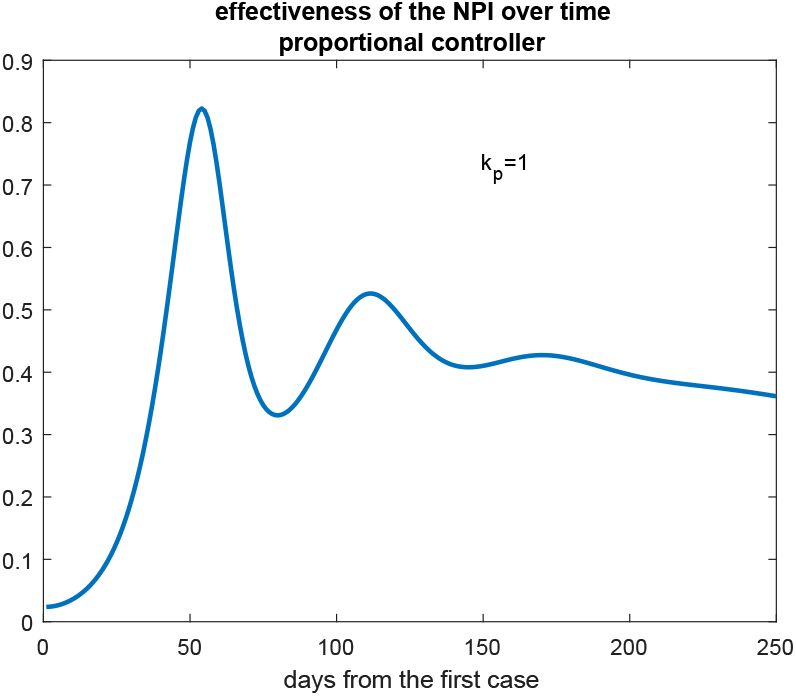
Control signal over time using a proportional controller.

The control signal presents a maximum value of 0.8227, and the area under the curve of the control signal vs. time is 98.8829. Of course, the smaller this action, the less the damage to the population and to the economy. The constant control signal equal to 0.4 presents an area under the curve equal to 100 and equal to 88.8 when it is applied after 4 weeks (see Table 1).

We must bear in mind that NPIs are determined by governmental or popular decisions, and hardly can change every day as the control signal calculated by the proportional controller does. Thus, we consider the application of NPIs with effectiveness shown in Fig. 9. The amplitudes and times of this control signal were obtained from that shown in Fig. 8. The detail of the trajectories of the states presented in Fig. 10 shows that there are no significant changes in the results presented. The maximum number of hospitalized people is 0.0057, the final number of deaths is 0.0132, and the area under the curve *u* vs. time is 87.38.

**Figure 9:**
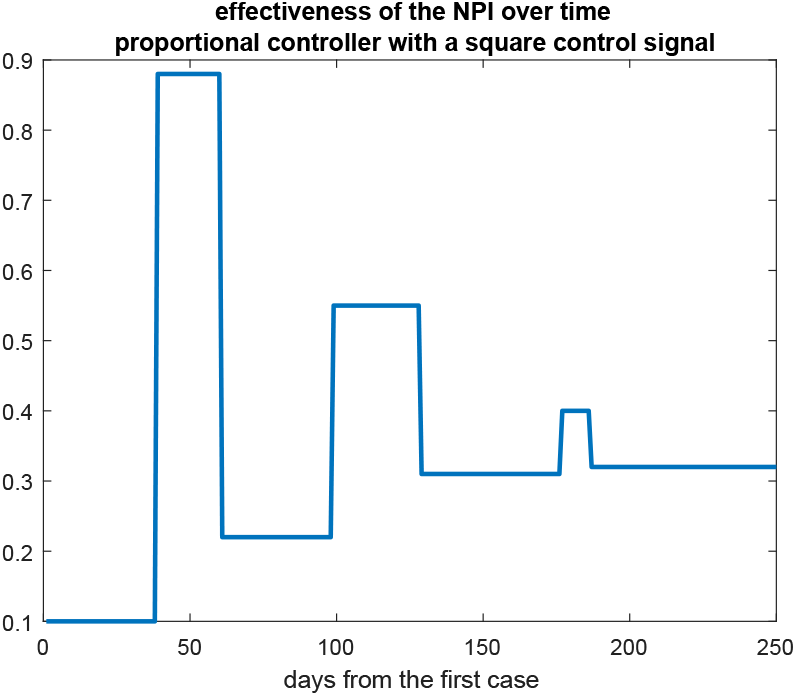
Step-shaped control signal over time using a proportional controller.

**Figure 10:**
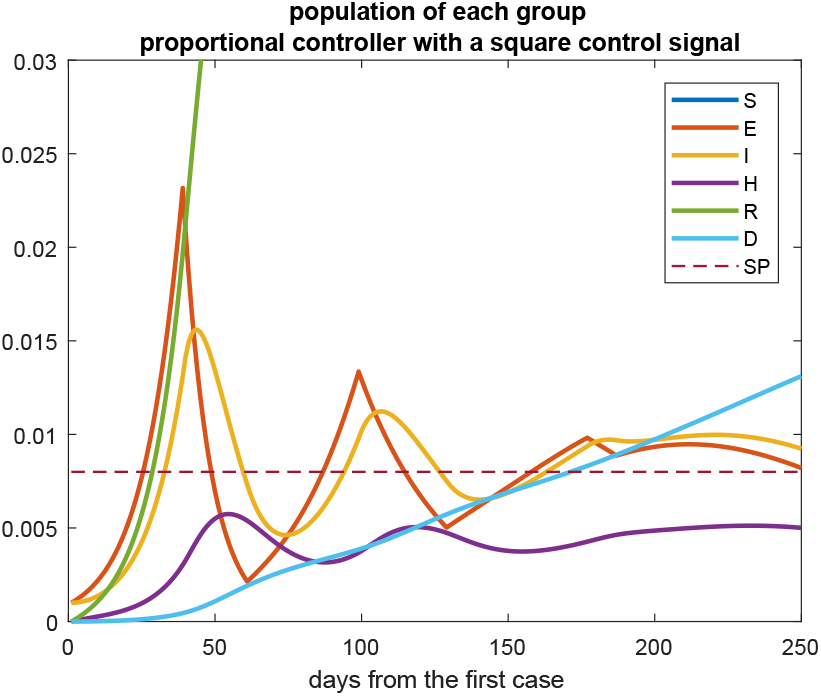
Detail of the trajectories of each population using a step-shaped control signal.

**Table 2:**
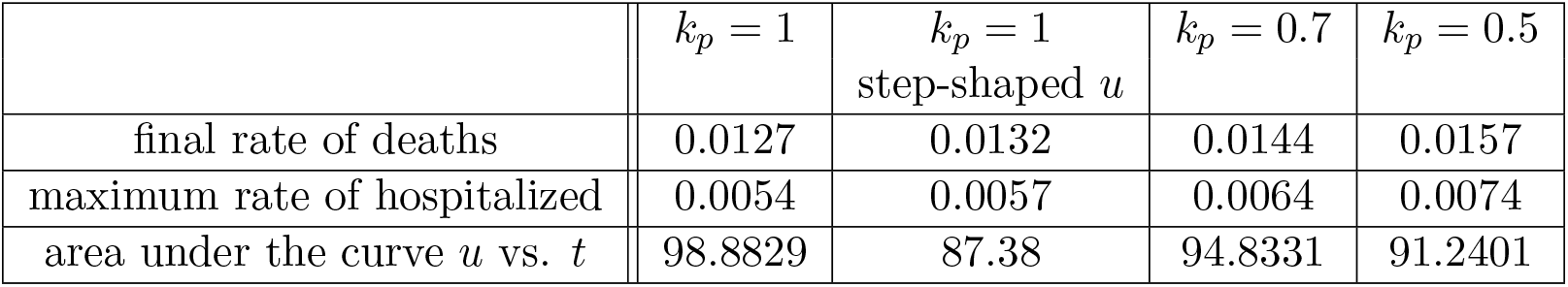
Main results of several proportional NPI on the SEIHRD model after 250 days, *SP* = 0.008.

Table 2 shows the main results of the application of NPIs calculated using a proportional controller with different values of the scalar gain *k_p_*.

## 3.3 Simulations with uncertain parameters and considering some noncompliance of the nonpharmaceutical interventions

In this section we consider the more realistic situation in which the parameters are partially unknown. As mentioned in Sec. 2 there are large uncertainties in the parameters, they diverge a lot according to the researched references and they are very different according to the country studied.

In this series of experiments, the parameter *α* is randomly chosen is also randomly chosen between 0.15 and 0.6. The parameter *β* between 0.008 and 0.04. The incubation time *γ*^−1^ between 2 and 6 days. The probability to present symptoms *p*_1_ between 40% and 80%. The time of recovering between 14 and 16 days, for both symptomatic or asymptomatic people. The probability to be hospitalized *p*_2_ is considered as a Gaussian distribution function of mean 0.19 and standard deviation of 0.1. The time to be hospitalized *δ*^−1^ is chosen between 3 and 7 days. The probability to die *p*_3_ between 10% and 16%. The time to die ∊^−1^ between 3 and 12 days. Finally, the time of recovering from hospitalization *μ*^−1^ is randomly chosen between 10 and 20 days.

In addition, we also consider that there exists some noncompliance of the nonpharmaceutical interventions. Hence, we apply to the system (1) a control signal with Gaussian distribution of mean 80% of that calculated in (3) with standard deviation of 10%, that is, we assume that there is 20% on average of noncompliance with the public measures adopted.

The initial conditions are also *I* = *E* = 0.001 and the gain is *k_p_* = 1. Fig. 11 shows the trajectories of the states of the model (1) during 250 days since the first symptomatic case arose.

**Figure 11:**
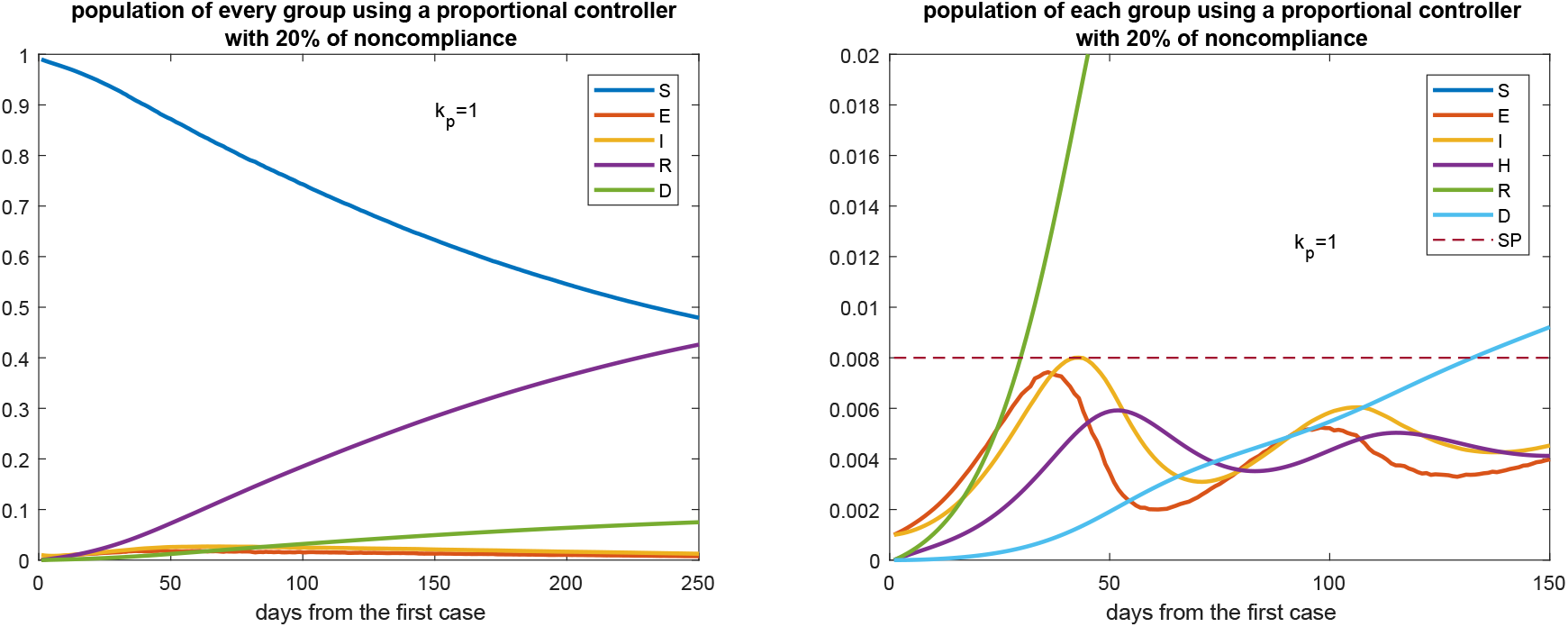
Evolution of every group over time with a proportional control action with gain *k_p_* = 1 (left) and considering 20% of noncompliance of the NPIs policies on average. The picture at the right is a zoom of that at the left. Set point equal to 0.008.

Fig. 12 shows the control signal vs. time.

**Figure 12:**
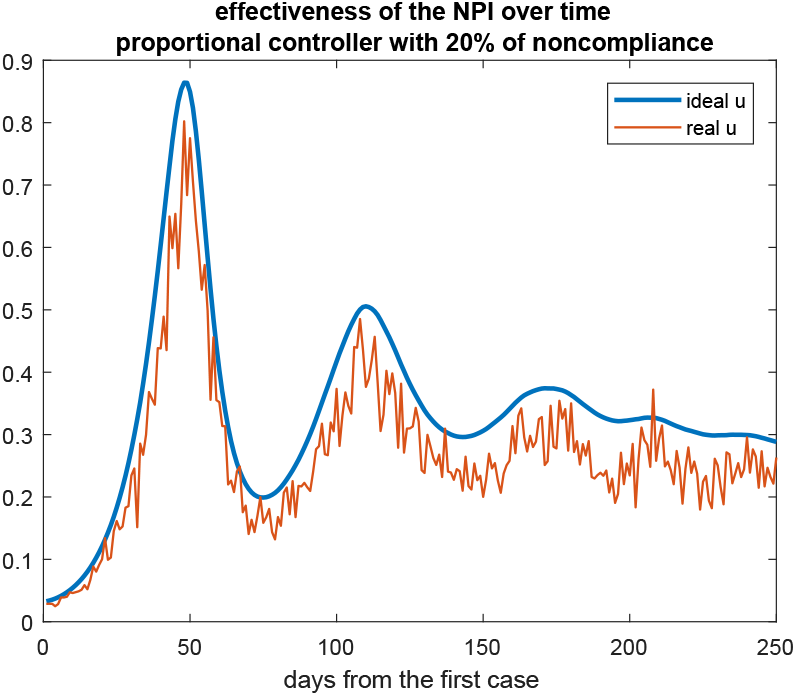
Control signal over time using a proportional controller and considering 20% of noncompliance of the NPIs policies on average. The blue curve is that calculated in (3), the red curve is the control signal considering the randomic noncompliance.

Table 3 reports some results extracted from this series of simulations.

The similarity of the results reported in Tables 2 and 3, as well as the trajectories shown in Figs. 7 and 11, show that the proportional controller is robust to parameter uncertainties and to some noncomplaince of the NPIs, which, of course, always occurs in practice.

**Table 3:** Main results of several proportional NPI on the SEIHRD model after 250 days considering 20% of noncompliance of the NPI on average, *SP* = 0.008.

| | $k_p = 1$ | $k_p = 0.7$ | $k_p = 0.5$ |
| --- | --- | --- | --- |
| final rate of deaths | 0.0225 | 0.0248 | 0.0735 |
| maximum rate of hospitalized | 0.0067 | 0.0076 | 0.0508 |
| area under the curve $u$ vs. $t$ | 73.7867 | 70.8435 | 77.4710 |

## 4 Conclusions

The proportional controller proposed to guide the adoption of NPIs shown its efficiency to keep the number of hospitalized people below a set point given by the health system capacity. Moreover, this very simple strategy is robust to parameters uncertainties and to some level of noncompliance of the public measures.

The control signal calculated by this method aims to guide the adoption of NPIs in order of minimizing the social impact and the economical damages.

As an example, recently the Argentine government relaxed some restrictions adopted in the quarantine period, allowing more economic and recreational activities in some cities. The only criterion used to adopt this measure was the number of days in which the number of infected people doubled (the so-called doubling time). Even thought this decision also can be considered as a closed loop control action, the criterion adopted is a little improvised.

An open question is how to translate the rate of effectiveness of the NPI calculated by the controller into concrete actions adopted by governments or public health authorities. Moreover, we must bear in mind that these measures cannot be continuously varied along the time, as the control signal does, but they are decisions that will remain valid for at least few days. However, although this issue is out of the scope of this paper, some decision can be changed every day, for example, the number of individuals with permission to leave their homes or the number of people allowed to get into a store, among other little decisions that can change every day according to the control variable suggests.

## Data Availability

all the data used in the article are available in https://www.worldometers.info/coronavirus/

## Footnotes

1 In Argentina, the daily death rate is 2.055 10^−5^.

2 At the moment, in the Covid-19 disease is an open question if a recovered person can get re-infected. Even though some cases were recently reported, the reinfection rate value appears to be statistically negligible based on early evidence.

3 This probability is the most difficult to determine. According to [Low20] up to 80% of the cases could be asymptomatic.

